# Specifications of the ACMG/AMP variant curation guidelines for the analysis of germline *ATM* sequence variants

**DOI:** 10.1101/2024.05.28.24307502

**Authors:** Marcy E. Richardson, Megan Holdren, Terra Brannan, Miguel de la Hoya, Amanda B. Spurdle, Sean V. Tavtigian, Colin C. Young, Lauren Zec, Susan Hiraki, Michael J. Anderson, Logan C. Walker, Shannon McNulty, Clare Turnbull, Marc Tischkowitz, Katherine Schon, Thomas Slavin, William D. Foulkes, Melissa Cline, Alvaro N. Monteiro, Tina Pesaran, Fergus J. Couch

**Affiliations:** Ambry Genetics, Aliso Viejo, CA, USA; Department of Laboratory Medicine and Pathology, Mayo Clinic, Rochester, MN, USA; Molecular Oncology Laboratory, Hospital Clínico San Carlos, IdISSC, 28040 Madrid, Spain; Population Health, QIMR Berghofer Medical Research Institute, Brisbane, QLD 4006, Australia; Department of Oncological Sciences and Huntsman Cancer Institute, University of Utah, Salt Lake City, UT, USA; Natera, Inc, San Carlos, CA, USA; GeneDx, Gaithersburg, MD, USA; Invitae Corporation, San Francisco, CA, USA; Department of Pathology and Biomedical Science, University of Otago, Christchurch, New Zealand; Department of Pathology and Laboratory Medicine, The University of North Carolina at Chapel Hill, Chapel Hill, NC, USA; Division of Genetics and Epidemiology, Institute of Cancer Research, London, UK; City of Hope Comprehensive Cancer Center, Duarte, CA, USA; Departments of Human Genetics, McGill University, Montreal, Quebec, Canada; UC Santa Cruz Genomics Institute, Mail Stop: Genomics, University of California, Santa Cruz, CA, USA; Department of Cancer Epidemiology, H Lee Moffitt Cancer Center & Research Institute, Tampa, FL, USA

## Abstract

The ClinGen Hereditary Breast, Ovarian and Pancreatic Cancer (HBOP) Variant Curation Expert Panel (VCEP) is composed of internationally recognized experts in clinical genetics, molecular biology and variant interpretation. This VCEP made specifications for ACMG/AMP guidelines for the ataxia telangiectasia mutated (*ATM*) gene according to the Food and Drug Administration (FDA)-approved ClinGen protocol. These gene-specific rules for *ATM* were modified from the American College of Medical Genetics and Association for Molecular Pathology (ACMG/AMP) guidelines and were tested against 33 *ATM* variants of various types and classifications in a pilot curation phase. The pilot revealed a majority agreement between the HBOP VCEP classifications and the ClinVar-deposited classifications. Six pilot variants had conflicting interpretations in ClinVar and reevaluation with the VCEP’s *ATM*-specific rules resulted in four that were classified as benign, one as likely pathogenic and one as a variant of uncertain significance (VUS) by the VCEP, improving the certainty of interpretations in the public domain. Overall, 28 the 33 pilot variants were not VUS leading to an 85% classification rate. The ClinGen-approved, modified rules demonstrated value for improved interpretation of variants in *ATM*.

## Introduction

The widespread adoption of low cost, high-throughput, next generation sequencing (NGS)-based multi-gene panel tests has led to a substantial increase in the detection of germline sequence variants. In 2015, in response to this increase, the American College of Medical Genetics and Genomics and the Association for Molecular Pathology (ACMG/AMP) provided a substantial update to their variant interpretation guidelines addressing many of the new challenges for variant interpretation ^1,2^. Because these guidelines are intended for use with any Mendelian disorder, gene- and disease-specific modifications may be needed to develop a tailored approach to variant classification. The process of tailoring variant interpretation guidelines is overseen by the National Institute of Health-funded Clinical Genome Resource (ClinGen) whose mission it is to develop an authoritative, comprehensive, central resource for expert-guided, gene- and variant-level information^3–5^. As part of this ClinGen initiative, the Hereditary Breast, Ovarian and Pancreatic Cancer (HBOP) Variant Curation Expert Panel (VCEP) formed in 2018, with a goal of specifying criteria of the 2015 ACMG/AMP baseline guidelines for clinical classification of variants in *ATM* (MIM 607585), *BARD1* (MIM 601593), *BRIP1* (MIM 605882), *CHEK2* (MIM 604373), *PALB2* (MIM 610355), *RAD51C* (MIM 602774), and *RAD51D* (MIM 602954) (https://clinicalgenome.org/affiliation/50039/). Based on the large number of variants and VUS in ClinVar, the Ataxia Telangiectasia mutated (*ATM*) tumor suppressor gene was selected for initial work of this VCEP.

*ATM* encodes a serine-threonine kinase involved in the cellular response to DNA damage^6^. Heterozygous loss-of-function (LoF) variants in *ATM* are associated with approximately 2-fold increased lifetime risks for breast cancer (MIM#114480) with a penetrance of 20-30%; and a 6.5-fold increased risk for pancreatic cancer ^7–11^. Biallelic pathogenic variants in *ATM* lead to the autosomal recessive disease Ataxia Telangiectasia (A-T) (MIM# 208900)], a severe, early-onset disorder characterized by progressive cerebellar ataxia and ocular telangiectasias ^12^ and increased cancer risk most commonly for leukemia and lymphomas ^13^. Large epidemiological and molecular studies have demonstrated that variants that cause A-T in the biallelic state are also expected to cause increased risk of breast and pancreatic cancer ^14,15^. As such, variants that cause A-T in the biallelic state are also considered by the HBOP VCEP to cause increased risk of breast and pancreatic cancer in the heterozygous state. Given the demonstrated increased risk for autosomal dominant and recessive disease, individuals with likely pathogenic/pathogenic (LP/P) variants in *ATM* may elect to increase cancer surveillance and/or be counseled for family planning. However, there are currently over 7,500 variants of uncertain significance (VUS) deposited to ClinVar, many of which are missense and non-coding variants (https://www.ncbi.nlm.nih.gov/clinvar/?term=atm%5Bgene%5D&redir=gene accessed 3/19/2024).

Therefore, the HBOP VCEP selected *ATM* for development of a validated set of variant classification rule specifications modeled on the baseline 2015 ACMG/AMP guidelines. The gene-specific rules for *ATM* along with application of these rules to curation of a series of *ATM* variants are described herein.

## Methods

### ClinGen HBOP VCEP

The HBOP VCEP formed in 2018 and is comprised of an international team of experts with relevant backgrounds in basic science research including protein functional analysis, clinical genetics, tumor pathology, computational principles, and/or variant interpretation. All members declared conflicts of interest as required by the FDA-approved ClinGen process, including several members who are full time employees at clinical diagnostic laboratories. The HBOP VCEP convened bi-weekly to consider the applicability, weight modifications, and gene-specific nuances of each of the categorical ACMG/AMP guidelines for *ATM*^1^. Initial rules were drafted based on evidence in the literature, internal laboratory data, and expert opinion and approved for pilot phase by ClinGen’s Sequence Variant Interpretation (SVI) group, who oversees this process.

### Pilot Phase

The *ATM*-specific rules were applied in a pilot test of 33 variants comprised of multiple different types (frameshift, nonsense, synonymous, intronic, canonical intronic, missense and structural variants), with different applicable evidence (high frequency variants, rare variants, variants identified in patients with A-T, variants in different functional domains, and variants tested in published functional assays), and/or selected for a variety of clinical assertions in ClinVar. Relevant clinical and allelic data from unpublished sources were solicited from the membership ahead of curation. Two curators independently evaluated variants and compared results. Differences were resolved first by discussion and agreement in a separate biocurator working group, that convened monthly. Differences were then escalated for a secondary review and consensus from the whole HBOP VCEP by vote. If needed, rules were modified or clarified in response to this process.

### Final ATM Rules

Modifications made in response to the pilot study were submitted to the ClinGen SVI for review. The final round of modifications, as recommended by the SVI, were implemented, and resubmitted for approval. Final interpretations for each of the pilot variants were curated into the Variant Curation Interface (VCI) and ultimately deposited to ClinVar. Classifications followed the original five-tier model (Benign, Likely Benign, Variant of Uncertain Significance, Likely Pathogenic and Pathogenic) and evidence combinations with a few modifications that are supported by a Bayesian framework^16^. The most recent *ATM* guidelines can be found on the Criteria Specification Registry and will be updated periodically as the HBOP VCEP continues their work (https://cspec.genome.network/cspec/ui/svi/doc/GN020).

## Results

### Rules not adopted for *ATM* by the HBOP VCEP (PS2, PS4_Moderate, PM1, PM6, PP1, PP2, PP4, PP5, BS2, BS4, BP1, BP3, BP5, BP6)

The HBOP VCEP chose not to adopt numerous ACMG/AMP codes for *ATM* for several reasons (Table 1). First, breast cancer is relatively common and the majority of it is non-hereditary, or sporadic. Second, hereditary and sporadic breast cancer cannot be distinguished from each other at this time. And third, ATM has low penetrance for breast cancer, conveying only two-fold risk which leads to substantial phenocopy and unaffected carriers of pathogenic variants within a family. The codes that were not adopted are detailed below.

**TABLE 1.**
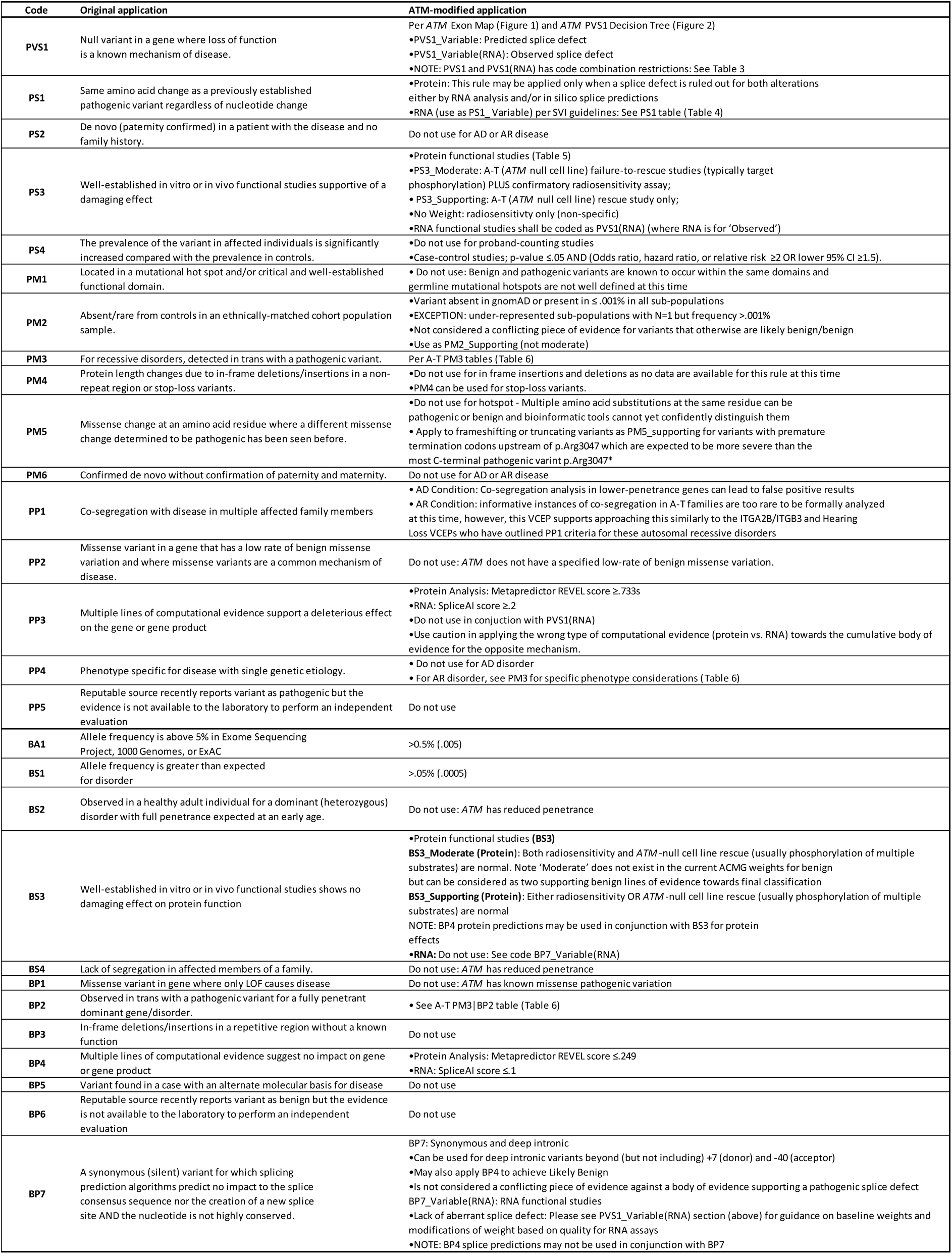
SUMMARY OF *ATM*-SPECIFIC RULES SPECIFICATIONS.

#### PS2/PM6: De novo

The observation of a *de novo* variant in the setting of a new disease is evidence towards pathogenicity. The use of *de novo* instances is not informative for *ATM* because breast cancer as a ‘new disease’ cannot be confidently established given the commonness of sporadic breast cancer.

#### PS4_Moderate: Proband Counting

With rare variants where a case control analysis cannot be statistically powered, an approximation called ‘proband counting’ can be used instead. In this method, affected probands can be weighted/counted towards pathogenicity once they reach a certain number that is designed to accommodate the disease and penetrance. It is most useful for pathognomonic gene-disease relationships with high penetrance. Because many genes cause breast cancer predisposition, and because penetrance is low, proband counting does not apply to *ATM*.

#### PP1/BS4: Co-segregation

Segregation of a disease and a variant within the same family is evidence for pathogenicity. However, genes conferring lower risk (Relative Risk = 2) for autosomal dominant conditions, should not be considered for co-segregation analysis because an unrealistic number of pedigrees is needed to obtain a true positive result whilst avoiding a false positive result. For example, in a gene with a Relative Risk of 2, like *ATM*, the probability of obtaining a true positive result for PP1 (as supporting strength) caps at 80% with 40 pedigrees, however that same circumstance also comes with a ∼3.5% chance of obtaining a false positive BP4 result ^17^. Regarding the use of BS4 (lack-of-segregation) in families with biallelic A-T is theoretically feasible, however, siblings with the same two variants as the A-T affected proband would be captured under the BP2 code which is a biallelic-unaffected patient.

#### PP4/BS2: Phenotype

A patient who has a phenotype that is highly specific for a disease or an unaffected patient who has not manifested disease can be used in the pathogenic (PP4) or benign (BS2) direction, respectively. However, since hereditary and sporadic breast and pancreatic cancer cannot be distinguished and because the penetrance is low for breast cancer, neither situation can be satisfied for *ATM*.

#### BP5: Variant present in a patient with an alternate mechanism for disease

This rule does not apply, because there are numerous examples of patients carrying both an *ATM* LP/P variant in addition to a second LP/P in other genes whose phenotype is not different than if they were carriers of a single pathogenic variant^18^.

#### PM1: Variant in a functional domain without benign variation

Although ATM has well established functional domains, there are many benign variants described in these domains, based on allele frequency and homozygous occurrences alone (https://gnomad.broadinstitute.org/gene/ENSG00000149311?dataset=gnomad_r2_1 accessed 3/19/2024).

#### PP2 and BP1: Low Rate of Benign Missense Variation

Pathogenic missense variants in *ATM* have been described, and there is not a specified low rate of benign missense variation.

#### BP3: in-frame indels in a repetitive region

There are insufficient data to support the use of in-frame deletions/insertions in a repetitive region without a known function.

#### PP5 and BP6: Reputable Source

These rules regarding reputable sources have been discontinued at the recommendation of the SVI ^19^

### Population Based Rules (BA1, BS1 and PM2_Supporting)

#### BA1 and BS1

The HBOP VCEP compared parameters for both the autosomal dominant and autosomal recessive conditions ascribed to *ATM* to estimate population allele frequency thresholds using the Whiffen/Ware calculator ^20^; https://cardiodb.org/allelefrequencyapp/). Because LP/P variants in *ATM* are considered a relatively infrequent cause of hereditary breast cancer the genetic heterogeneity was set to 0.02: in other words, as if 2% of hereditary breast cancer cases are caused by *ATM* LP/P variants. The allelic heterogeneity was conservatively set to 1.0: in other words, assuming that there is only one LP/P variant that causes *ATM*-related breast cancers. Lastly, the penetrance for *ATM* and breast cancer was conservatively set to 0.2 based on data from multiple studies of hereditary breast cancer ^10,11^. Using these parameters, and a prevalence of 1:8 women for breast cancer, the maximum credible allele frequency was 0.625%. Similarly, for A-T the autosomal recessive inheritance was selected along with a prevalence of 1:40,000 ^21–25^. As *ATM* is the only gene that causes A-T, the genetic heterogeneity was set to 1.0 and penetrance was set to 0.90. Using these parameters, the maximum credible allele frequency is 0.527%. Given the conservative parameters put into the calculator and to simplify, the BA1 threshold was set to 0.5%. For BS1, all parameters remained the same except for the extremely conservative allelic heterogeneity value, which was dropped to 0.10, leading to an order-of-magnitude decrease in the maximum credible allele frequency of 0.05%. In applying these frequency codes, statistical models should be considered to account for error related to sample size such as the filtering allele frequency in gnomAD ^26^.

#### PM2

ClinGen has deviated from the Richards *et al* ACMG/AMP guidelines for PM2 and now recommends that this evidence code be uniformly down weighted to PM2_Supporting (https://www.clinicalgenome.org/site/assets/files/5182/pm2_-_svi_recommendation_-_approved_sept2020.pdf). This recommendation was adopted for rare *ATM* variants. Due to the incomplete penetrance, it is reasonable to expect that unaffected carriers are present in the general population. As such a variant does not need to be absent in the general population to apply PM2_Supporting. For *ATM*, rarity is considered as a general population frequency of ≤0.001% in each subpopulation. Any alteration that exceeds 0.001% in a large general population database but for which there is only one carrier is still considered eligible for PM2_Supporting.

### Loss-of-Function Codes (PVS1 and PVS1(RNA))

#### PVS1

LoF is the mechanism of disease for *ATM* ^22,27,28^. The rules governing the application and appropriate weight of PVS1 are based on the ClinGen SVI recommendations ^29^. There are five variant types that fall under the PVS1 category: nonsense and frameshift alterations; canonical (+/-1,2) splice site alterations (and some last-nucleotide alterations), gross deletions, gross duplications, and initiation codon alterations. Several features influence the weight ascribed to PVS1 including: 1) nonsense-mediated RNA decay (NMD); 2) the impact of an NMD-escaping effect on a critical functional domain 3) the size of the NMD-escaping effect relative to the size of the protein; and 4) gene-specific features.

##### *ATM* canonical transcript

The reference transcripts considered by this VCEP are NM_000051.3/ENST00000278616.8. All exons from this transcript are considered constitutive exons without major alternative splice isoforms that could result in a rescue of PVS1-eligible variants ^30–33^. This transcript contains a non-coding first exon (Exon 1) and 62 subsequent coding exons: Exon 2-63 (Figure 1). Of note, *ATM* may be annotated with four non-coding first exons leading to legacy nomenclature references in historical data.

**Figure 1.**
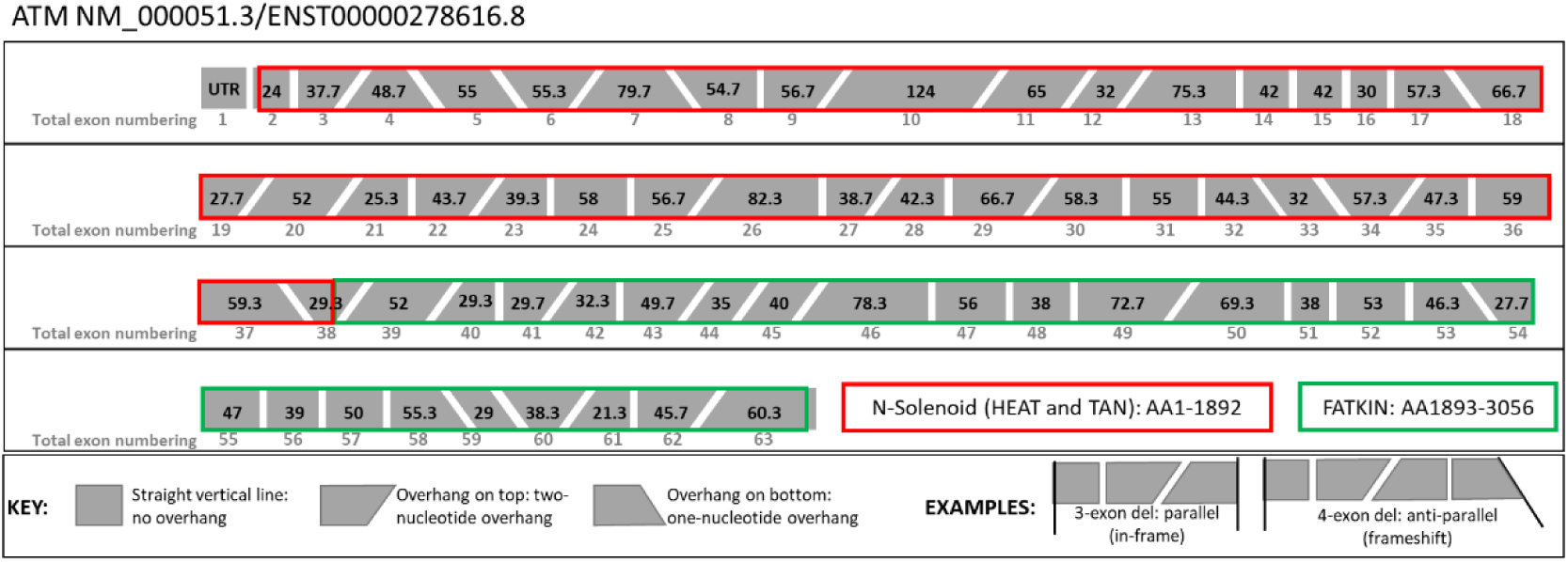
ATM Exon Numbering and Reading Frame. The ATM gene is depicted exon-by-exon. The amino acid size of each exon is depicted within the boxes in black text. The two major functional domains are outlined in red (N-Solenoid, comprised of sub-domains HEAT Repeat and TAN domain) and green outline (FATKIN domain comprised of the FAT and FAT-C sub-domains). Each exon is shaped to indicate the number of overhanging nucleotides at either end which will assist in determining any reading-frame changes from gross deletions or duplications of whole single- or multi-exons. A vertical line indicates a blunt start or end with no overhanging nucleotides. An upper overhang on either side represents a two-nucleotide overhang; A lower overhang represents a single-nucleotide overhang on that side. To use this diagram, a line drawn at the start and end of a deletion or duplication will be either parallel (in-frame event) or non-parallel (frameshift) as in the examples.

##### *ATM* Functional Domains

*ATM* is comprised of two main functional domains: the N-Solenoid domain and the FATKIN domain. The N-terminal half of the protein is an α-solenoid structure (N-Solenoid) (amino acids 1-1892)^34^ that is able to interact with nucleic acids and various protein partners. The **P**hospho**i**nositide **3**-**K**inase domain (PI3-K), the **F**ocal **A**dhesion **T**argeting (FAT), and the **F**ocal **A**dhesion **T**argeting **C**arboxyterminal (FATC) collectively comprise the FATKIN domain of ATM (Figure 1). The FATKIN domain is directly responsible for ATM kinase function, which is essential for tumor suppressor activity. Therefore, the FATKIN domain is considered critical for protein function and NMD-escaping variants, including in frame losses and truncations between p.Leu2980 and p.Arg3047, that adversely affect the FATKIN domain are given PVS1 as Very_Strong ^35–38^. The N-Solenoid domain is thought to be important for protein function because there are numerous patients affected with A-T who carry alterations that are known to lead to in-frame losses in the N-Solenoid domain (Supplementary Figure 1) ^30,32,39–49^. However, compared to the FATKIN domain, there are relatively few missense pathogenic mutations (https://www.ncbi.nlm.nih.gov/clinvar/?term=atm%5Bgene%5D&redir=gene accessed 3/19/2024). Because of this, in-frame single- or multi-exon losses impacting the N-Solenoid domain can receive PVS1_Strong.

##### PVS1 Eligibility Boundaries

Because pathogenic variants in *ATM* cause both A-T in a biallelic state and cancer predisposition in a heterozygous state, this VCEP was able to leverage evidence from A-T cohorts to inform PVS1 boundaries. At the N-terminus, it was determined that variants destroying the initiation codon are ascribed PVS1 as Very_Strong due to the identification of numerous A-T affected individuals harboring p.Met1? variants ^45,47,50–53^. In addition, LoF alterations lying between p.Met1? and the next downstream, in-frame methionine at p.Met94 have also been observed in A-T patients supporting that downstream methionine residues are unable to serve as an alternate start codon that would produce a rescue effect ^54–56^. At the C-terminus, p.Arg3047 is considered the last critical amino acid based on many reports of a nonsense variant at this position in patients with A-T ^30,43,45,47,57–62^. Therefore, LoF alterations impacting codons between p.Met1 and p.Arg3047 are eligible for PVS1 at varying weights according to the PVS1 Decision Tree (Figure 2).

**Figure 2.**
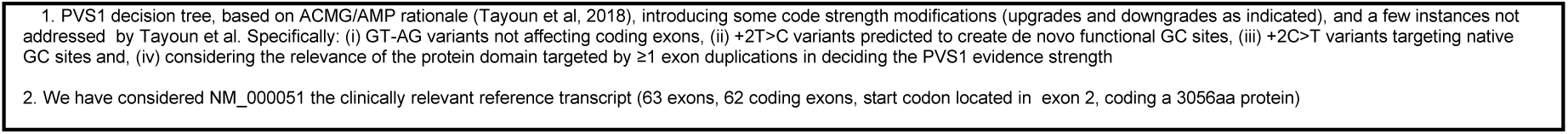
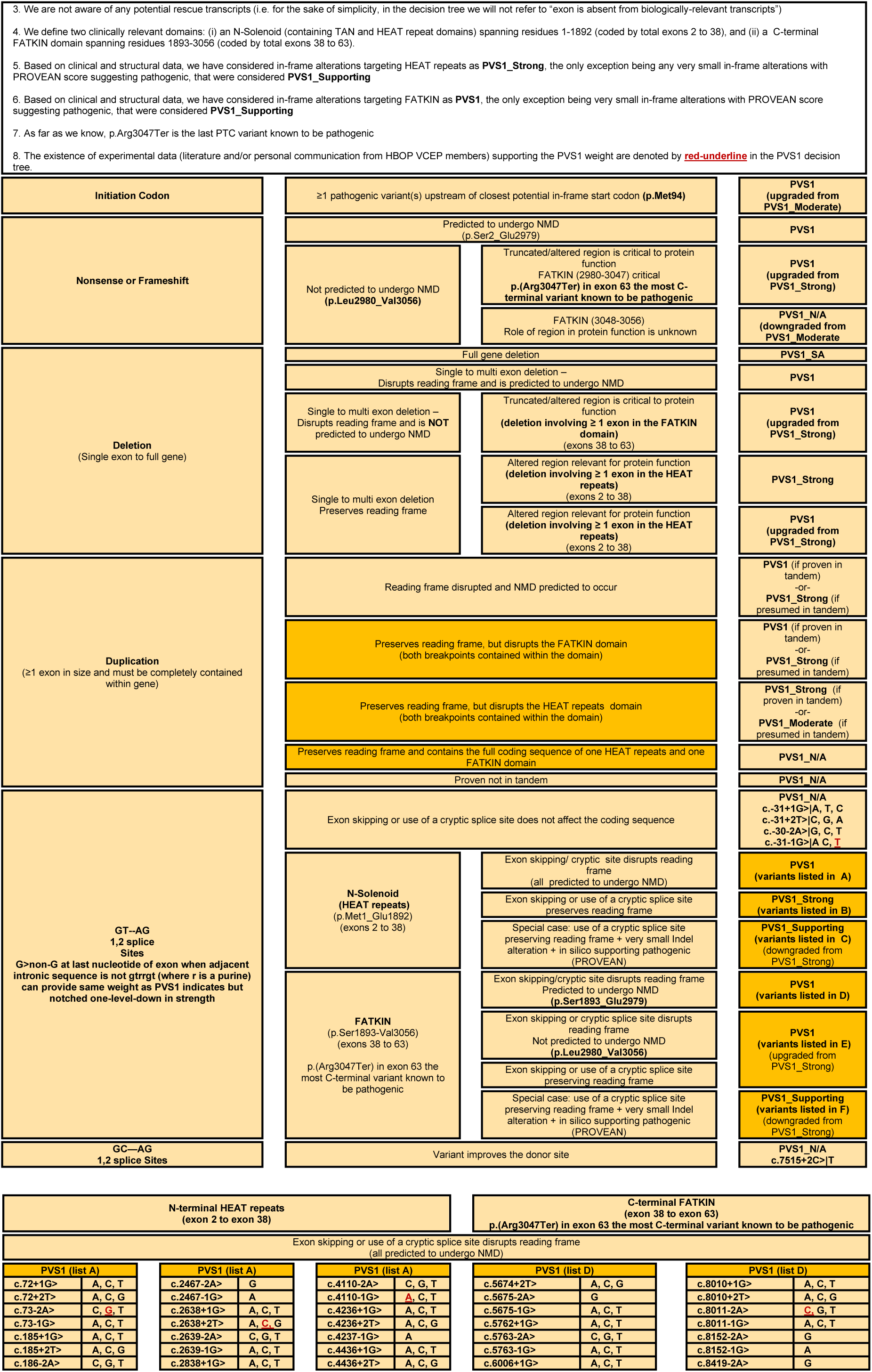
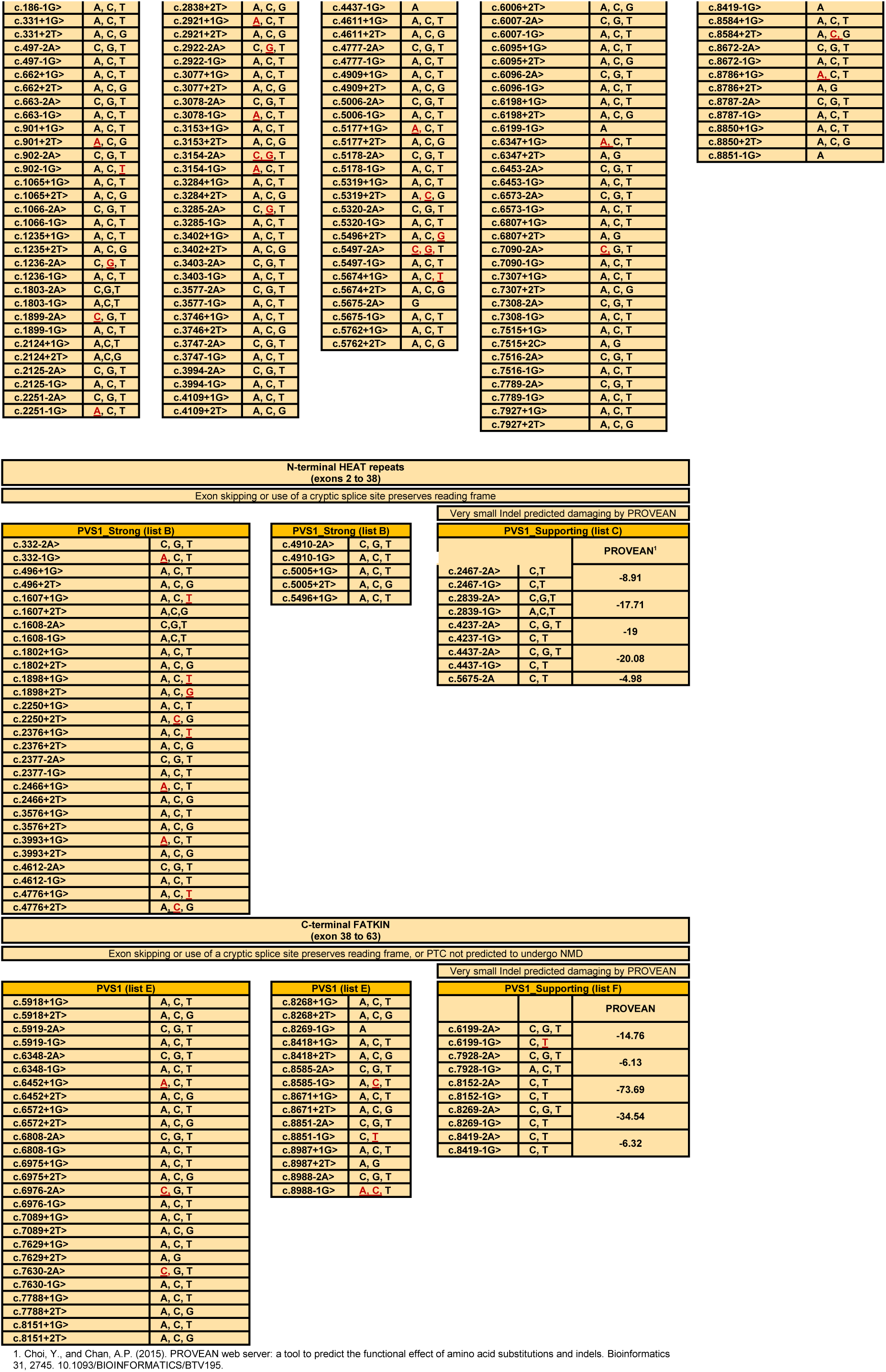
PVS1 Decision Tree for ATM. PVS1 eligible variant types are split into five categories: initiation codon variants, nonsense and frameshift variants, ≥1 exon deletions, ≥1 exon duplications and last NT/canonical splice variants. Considerations related to NMD, N and C terminal boundaries, domain involvement, tandemness, and splice prediction/observation inform the weight that can be afforded to the PVS1 criterion. Small in-frame events were predicted with PROVEAN and scores are provided. Nucleotides in red-underline have splice effects reported in the literature.

##### Gross deletions

Single-to-multi-exon deletions that are frameshifting and NMD-prone receive PVS1 weight at Very_Strong as per the original 2018 PVS1 guidelines^29^. Alterations producing NMD-escaping transcripts that adversely affect the N-Solenoid receive PVS1_Strong and those adversely affecting the FATKIN domain receive PVS1 as Very_Strong. The HBOP VCEP has made a diagram to assist with discerning the reading frame disruption of gross deletions and duplications (Figure 1).

##### Gross duplications

Single-to-multi-exon duplications that do not involve either the 5’ or 3’ untranslated regions (UTRs) are eligible for PVS1 weight whether they are confirmed or presumed in tandem. PVS1 (as Very_Strong) and PVS1_Strong can be applied for in-frame events confirmed or presumed to disrupt the FATKIN domain, respectively; and PVS1_Strong and PVS1_Moderate can be applied for in-frame events confirmed or presumed to disrupt the N-Solenoid domain, respectively. Care should be taken to ensure that the functional domains are *disrupted* by the duplication which means both the 5’ and 3’ breakpoint of the duplication must be within the same domain. Duplications that have one breakpoint in the N-solenoid domain and one breakpoint in the FATKIN domain do not disrupt either domain and do not receive any PVS1 weight.

##### Splice variants

Canonical splice variants are defined as the +/-1 and 2 positions in the introns surrounding an exon as well as some alterations at the last nucleotide of the exon. If the sequence does not conform to the consensus U2 donor site of Ggtrrgt (where the capital G is the last-nucleotide position of the exon and where r is any purine) then the impact of a last nucleotide substitution on splicing is expected to be greater. Such alterations are eligible for PVS1 weight but are reduced by one strength level from the corresponding +1,2 baseline weight provided in the PVS1 Decision Tree (Figure 2). Each possible +/-1,2 splice variant is parsed into a PVS1 list (A to F) depending on reading frame and impact on the N-Solenoid or FATKIN domains (Figure 2). Figure 2 was informed by *in silico* score from SpliceAI and/or PROVEAN, in conjunction with published and unpublished splicing data. Of note, there are several variants that receive PVS1_Supporting because they are predicted to make use of an in-frame alternate splice site that preserves the reading frame and leads to a small insertion or deletion that is predicted by PROVEAN to be deleterious (Figure 2, Lists C and F). There are also several candidate variants that do not receive any PVS1 weight because they are +2T>C alterations that do not have a predicted splice impact by SpliceAI. Although rare, +2T>C alterations are known to produce predominantly wildtype transcripts ^63^. There is also one splice site at *ATM* c.7515+2 that is atypical in that it has a native cytosine instead of the consensus thymine. Therefore, a C>T substitution here is predicted to improve the native splice sequence and it receives no PVS1 weight.

##### PVS1(RNA)

Any spliceogenic variant, whether canonical, exonic, or deeper intronic, that is confirmed by RNA studies to have a deleterious splice defect can be coded as PVS1(RNA). The application of PVS1(RNA) supplants any other predictive lines of evidence (PVS1 or PP3). Of note, PS3, the code for functional data supporting a pathogenic event, is not used for RNA data because it is reserved for downstream (e.g. protein) functional effects which can be observed in conjunction with an RNA defect and applied in addition to PVS1(RNA). The weight for PVS1(RNA) can be variably ascribed based on curator judgement of the quality and quantitative result of the RNA assay according to recent recommendations by the SVI ^64^. In contrast, RNA functional studies establishing a lack of aberrant splicing can be coded as BP7(RNA). The weight for BP7(RNA) can be variably ascribed from Supporting to Strong based on curator judgment of the quality of the RNA assay.

### Computational/predictive data-driven rules (PS1, PM4, PM5, PP3, BP4, BP7)

#### PS1

A variant that produces the same protein change as a known pathogenic alteration can be given PS1 towards pathogenicity. This rule may only be applied when a splice defect is ruled out for both the known LP/P alteration and the variant under evaluation by *in silico* splice predictions or RNA evidence. If splicing is a factor for both variants, PS1 can be used as an RNA hotspot and the weight applied is per the ClinGen SVI recommendations (Table 2)^64^.

**TABLE 2.** PS1 CODE WEIGHTS FOR VARIANTS WITH SAME PREDICTED SPLICING EVENT AS KNOWN (LIKELY) PATHOGENIC VARIANT.

|  |  |  |  |  |
| --- | --- | --- | --- | --- |
| Variant under assessment (VUA) | Baseline computational/predictive code applicable to VUA | Position of comparison variant relative to VUA | PS1 code applicable to VUA |  |
|  |  |  | with P comparison variant | with LP comparison variant |
| Located outside splice donor/acceptor $\pm 1,2$ dinucleotide positions | PP3 | same nucleotide | PS1 | PS1_Moderate |
| | PP3 | within same splice donor/acceptor motif (including at $\pm 1,2$ positions) | PS1_Moderate | PS1_Supporting |
| Located at splice donor/acceptor $\pm 1,2$ dinucleotide positions | PVS1 | within same splice donor/acceptor $\pm 1,2$ dinucleotide | PS1_Supporting | N/A |
| | PVS1 | within same splice donor/acceptor region, but outside $\pm 1,2$ dinucleotide<sup>a</sup> | PS1_Supporting | PS1_Supporting |
| | PVS1_Strong, PVS1_Moderate, or PVS1_Supporting | within same splice donor/acceptor $\pm 1,2$ dinucleotide | PS1 | N/A |
| | PVS1_Strong, PVS1_Moderate, or PVS1_Supporting | within same splice donor/acceptor region, but outside $\pm 1,2$ dinucleotide<sup>a</sup> | PS1_Moderate | PS1_Supporting |
| Prerequisite for all: the predicted event of the VUA must precisely match the predicted event of the comparison (likely) pathogenic variant (e.g., both predicted to lead to exon skipping, or both to lead to enhanced use of a cryptic splice motif, AND the strength of the prediction for the VUA must be of similar or higher strength than the strength of the prediction for the comparison [likely] pathogenic variant). For an exonic variant, predicted or proven functional effect of missense substitution(s) encoded by the VUA and (likely) pathogenic variant should also be considered before application of this code. Dinucleotide positions refer to donor and acceptor dinucleotides in reference transcript(s) used for curation. Designated donor and acceptor motif ranges should be based on position weight matrices for intron category (see methods). For GT-AG introns these are defined as follows: the donor motif, last 3 bases of the exon and 6 nucleotides of intronic sequence adjacent to the exon; acceptor motif, first base of the exon and 20 nucleotides upstream from the exon boundary. Consider other motif ranges for non-GT-AG introns. |  |  |  |  |
| <sup>a</sup> If relevant, splicing assay data for a pathogenic variant outside a $\pm 1,2$ dinucleotide position may be used to update a PVS1 decision tree and hence the applicable PVS1 code for a $\pm 1,2$ dinucleotide variant. | | | | |

#### PM4

In frame deletions and insertions as well as variants disrupting the native stop codon may be eligible for PM4. However, for *ATM*, there are no data available at this time to inform the use of in-frame insertions or deletions. Stop-loss variants in *ATM* are eligible for PM4 due to the identification of numerous A-T patients harboring such pathogenic alterations ^45,56^.

#### PM5

This rule is ascribed to missense variants at an amino acid residue where another pathogenic missense alteration has been identified. However, amino acid substitutions at a single residue in *ATM* can be pathogenic or benign. Thus, the use of this rule is not recommended. However, this rule has been co-opted as PM5_Supporting to increase the evidence for pathogenicity for LoF alterations being ascribed PVS1 or PVS1(RNA) as Very_Strong. This rule is governed by *ATM*’s lack of alternative splicing events that would produce a functional protein leading to a putative rescue of LoF alterations by splicing the variant out. In this manner, the use of PVS1 and PM5_Supporting will classify all *ATM* LoF variants as likely pathogenic even if they do not meet PM2_Supporting. PVS1/PVS1(RNA)-eligible variants (applied as Very_Strong) that do meet rarity (PM2_Supporting) will be classified as likely pathogenic with the addition of PM5_Supporting.

#### PP3/BP4 Protein

This VCEP favors the use of the metapredictor REVEL for single nucleotide variation and Provean for small in-frame indels as a single predictor to anticipate the impact of a protein change ^65–67^. A REVEL score ≥0.733 is considered damaging (PP3). And a score ≤0.249 is considered neutral. This threshold is based on the general recommendation and not derived as an ATM-specific threshold at this time^65^. This was further supported by application to prediction of damaging effect in large functional datasets for multiple cancer genes^68^.

#### PP3/BP4 RNA

The VCEP uses SpliceAI as a sole predictor due to its ability to accurately predict loss of native splice sites and creation of cryptic sites ^69^. This VCEP did not declare gene-specific thresholds for SpliceAI but recommends those set forth by the SVI in applying PP3 to non-canonical splice variants with a SpliceAI score of ≥0.2 and BP4 to variants with a SpliceAI score ≤0.1 ^64^. In the event that RNA data are available and they reflect a substantial variant-specific impact, do not use both PVS1(RNA) and PP3 or BP4. However, in the event that RNA data are available and they reflect no variant-specific impacts, PP3 or BP4 may be applied in conjunction with BP7(RNA) (See Table 3). BP4 may also be used in conjunction with BP7 (see below)^64^.

**TABLE 3.** RESTRICTIONS ON COMBINING CRITERIA.

|  | PP3 | PS3\|BS3 | PS1 | PVS1 | PVS1 (RNA) | BP4 | BP7 | BP7 (RNA) |
| --- | --- | --- | --- | --- | --- | --- | --- | --- |
| PP3 |  | ✓ | ✓ | X | X | N/A | N/A | ✓ |
| PS3\|BS3 | ✓ |  | N/A | ✓ | ✓ | ✓ | ✓ | ✓ |
| PS1 | ✓ | N/A |  | N/A | N/A | N/A | N/A | N/A |
| PVS1 | X | ✓ | N/A |  | X | X | X | X |
| PVS1(RNA) | X | ✓ | N/A | X |  | X | X | X |
| BP4 | N/A | ✓ | N/A | X | X |  | ✓ | ✓ |
| BP7 | X | ✓ | N/A | X | X | ✓ |  | X |
| BP7(RNA) | ✓ | N/A | N/A | X | X | ✓ | X |  |

N/A: Not applicable because the codes are unrelated

#### BP7

This rule was originally intended for synonymous variants, however, the VCEP applied the rule to deep-intronic variants beyond (but not including) +7 at the donor site and −40 at the acceptor site. Per the SVI’s recent guidance, this code is to be applied only when BP4 is met, in which case both BP4 and BP7 would be applied ^64^ (See Table 3). Using these modifications, many synonymous and deep intronic variants can be classified towards benign by applying both BP7 and BP4, in the absence of conflicting data.

### Functional evidence (PS3/BS3)

#### PS3/BS3

This is applied to protein functional studies or studies that are downstream of RNA effects. For *ATM*, there are multiple well-established functional studies that employ the use of *ATM*-null cell lines to observe the general rescue of radiosensitivity and/or ATM-specific events such as phosphorylation of ATM substrates (Table 4) ^46,70,71^. Because many of the published assays have only a few variants, they contain insufficient known-pathogenic and known-benign controls for Bayesian validation^72^. However, because variant controls in several studies behave as expected in these assays the VCEP has approved a maximum weight of PS3_Moderate and BS3_Moderate for a combination of functional studies that are concordant for a non-functional or functional result, respectively. For non-functional results to be used as PS3_Moderate, both an ATM-specific functional result and a non-specific radiosensitivity functional result should agree. If there is disagreement between results then no weight should be applied towards PS3. If only the ATM-specific-study (e.g. ATM auto- or trans-phosphorylation at specific residues) result is available and reflects non-functional, a maximum weight of PS3_Supporting can be given. However, because a non-functional result from a radiosensitivity assay is not specific to an ATM defect, a non-functional result in a radiosensitivity assay alone does not achieve any PS3 weight. In the benign direction, a neutral result in either an ATM-specific assay or a radiosensitivity assay can be ascribed BS3_Supporting per each. Of note, both PP3/BP4 *in silico* protein predictions and PS3/BP4 protein functional studies can be co-applied.

**TABLE 4.** ATM FUNCTIONAL STUDIES.

| **ATM kinase activity** |  |  |  |
| --- | --- | --- | --- |
| **PMID** | 18634022 | 19431188 | 11805335 |
| **DOI / link** | 10.1002/humu.20805 | DOI: 10.1002/humu.21034 | 10.1073/pnas.012329699 |
| **Author** | Mitui | Barone | Scott |
| **Year** | 2009 | 2009 | 2002 |
| **Assay (general description)** | Stable transfection of ATM-based cDNA constructs in an ATM null cell line; ATM kinase activity assayed by Western Blotting of ATM substrates. | Stable transfection of ATM-based cDNA constructs in an ATM null cell line; ATM kinase activity assayed by Western Blotting of ATM substrates. | Stable transfection of ATM-based cDNA constructs in an ATM null cell line; ATM kinase activity assayed by Western Blotting of ATM substrates. |
| **Material used (patient cells, engineered variants, cell lines, animal model, etc.)** | ATM null patient LCL cells (AT7LA1-homozygous for a truncating mutation-c.1563_1564delAG); variants introduced by site-directed mutagenesis. | ATM null patient LCLs (patient 118-3 with two truncating mutations, c.796_797insGATT and c.2921+1G>A) ; variants introduced by site-directed mutagenesis. | ATM null patient LCLs (AT1ABR); variants introduced by site-directed mutagenesis. |
| **Readout type (qualitative/quantitative)** | qualitative- bands on a gel | quantitative- Western blot images scanned using a densitometer | qualitative-band on a gel |
| **Readout description** | phosphorylation status of ATM-S1981 and SMC1 (at S957 or S966) tested by Western Blot 1 hr after cells were irradiated with 2 or 10 Gy. Cells were treated with CdCl<sub>2</sub> to induce ATM expression from a metallothionein promoter II. | Phosphorylation status of SMC1 (Ser-966); NBS1 (Ser-343); CHK2 (Thr-68); p53 (Ser-15); ATM serine 1981 tested by Western Blot at multiple timepoints after cells were irradiated with 5 gray (0, 30 mins, 60 mins) or mock irradiated. Cells were treated with zinc chloride to induce ATM expression from a metallothionein promoter II. | Cells were treated with 6 Gy of ionizing radiation and kinase assays were performed 1 hr after radiation. Phosphorylation status of p53-Ser-15 was assayed directly on cell lysates ("in vivo") or by immunoprecipitation of flag-tagged ATM from cell extracts followed by phosphorylation of a p53 substrate (p531-40, "in vitro"). Cells were treated with CdCl<sub>2</sub> 6 h before irradiation to induce ATM expression from a metallothionein promoter II. |
| **Biological replicates (met/not met)** | uncertain/not described | not met | not met |
| **Technical replicates (met/not met); description** | uncertain/not described | Y; 3; not sure if biological or technical | uncertain/not described |
| **Basic positive control (met/not met); description** | met; WT cDNA (pMAT1) and ATM WT LCLs (NAT2) | met; WT cDNA (pTAM2) | met; WT cDNA (pMAT3) |
| **Basic negative control (met/not met); description** | met; ATM null cells (AT7LA1) | met; empty cDNA (pMEP4) | met; un-induced cells |
| **Validation controls P/LP (#)** | 0; c.5908C>T (p.Q1970*) - unclear if tested in this assay/data not shown | 0 | 5 AT mutants<br>"c.7636del9,p.SRI2564del3; c.8147T>C, p.V2716A; c.8546G>C p.R2849P; c.8599G>C p.G2867R; c.7987delGTT p.V2662del" |
| **Validation controls B/LB (#)** | 0; c.1744T>C(p.F582L) and c.2119T>C (p.S707P)- unclear if tested in this assay/data not shown | 0 | 0 |
| **Statistical analysis (general description)** | uncertain/not described | uncertain/not specified | uncertain/not described |
| **Threshold for normal readout** | authors label a normal readout as "normal" in table 1. "normal" is based on the output of the WT construct (pMAT1) | WT level of kinase activity (group 1) (relative to WT pTAM2) | not provided, but curator can distinguish as present or absent |
| **Threshold for abnormal readout** | authors label an abnormal read out as ND (non-detectable); TD, (trace detected) in table 1. | no detectable kinase activity (group 2) or reduced level of kinase activity (group 3), (relative to WT pTAM2). Authors note that ATM Serine 1981 phosphorylation may not be a good indicator of overall ATM kinase activity. | not provided, but curator can distinguish as present or absent |
| **Approved assay (y/n)** | Y | Y | Y |
| **Proposed strength** | **PS3_supporting**<br>**BS3_Supporting** | **PS3_Supporting;**<br>**BS3_supporting** | **PS3_Supporting**<br>**BS3_supporting** |
| **ATM radiosensitivity** |  |  |  |
| **PMID** | 18634022 | 11805335 | 11805335 |
| **DOI / link** | 10.1002/humu.20805 | 10.1073/pnas.012329699 | 10.1073/pnas.012329699 |
| **Author** | Mitui | Scott | Scott |
| **Year** | 2009 | 2002 | 2002 |
| **Assay (general description)** | Stable expression of cDNA constructs in an ATM null cell line; cellular radiosensitivity was assayed as a read-out for ATM function. | Stable transfection of ATM-based cDNA constructs in an ATM null cell line; cellular radiosensitivity was assayed as a read-out for ATM function. | Stable transfection of ATM-based cDNA constructs in an ATM null cell line; cellular radiosensitivity was assayed as a read-out for ATM function. |
| **Material used (patient cells, engineered variants, cell lines, animal model, etc.)** | ATM null patient LCL cells (AT7LA1-homozygous for a truncating mutation- c.1563_1564delAG); variants introduced by site-directed mutagenesis. | ATM null patient LCLs (AT1ABR); variants introduced by site-directed mutagenesis. | ATM null patient LCLs (AT1ABR); variants introduced by site-directed mutagenesis. |
| **Readout type (qualitative/quantitative)** | quantitative | quantitative | quantitative |
| **Readout description** | % survival fraction (SF) ( determined by MTT staining of viable cell colonies after irradiation with 1 gy ). Cells were treated with CdCl2 to induce ATM expression from a metallothionine promoter II. | Cells were treated with 1 Gy of y-rays and fifty metaphases were analyzed for each sample, and radiation induced chromosome aberrations (ICAs) were determined. Cells were treated with CdCl2 6 h before irradiation to induce ATM expression from a metallothionine promoter II | Cells were treated with 1-4 Gy of y-rays and the number of viable cells was determined daily up to 4 days post irradiation. Cells were treated with CdCl2 6 h before irradiation to induce ATM expression from a metallothionine promoter II. |
| **Biological replicates (met/not met)** | uncertain/not specified | uncertain/not specified |  |
| **Technical replicates (met/not met); description** | uncertain/not specified | uncertain/not specified | 3; unclear if biological or technical |
| **Basic positive control (met/not met); description** | met; WT cDNA (pMAT1) and ATM WT LCLs (NAT9) | met; WT cDNA and WT ATM cells (C3ABR) | met; WT cDNA and WT ATM cells (C3ABR) |
| **Basic negative control (met/not met); description** | met; ATM null cells (AT7LA1) | met; ATM null cells (AT1ABR) | met; ATM null cells (AT1ABR) |
| **Validation controls P/LP (#)** | 0; c.5908C>T (p.Q1970*) (not clear if tested in this assay/data not shown) | 5 AT mutants<br>"c.7636del9,p.SRI2564del3;<br>c.8147T>C, p.V2716A;<br>c.8546G>C p.R2849P;<br>c.8599G>C p.G2867R;<br>c.7987delGTT p.V2662del" | 5 AT mutants<br>"c.7636del9,p.SRI2564del3; c.8147T>C,<br>p.V2716A; c.8546G>C p.R2849P;<br>c.8599G>C p.G2867R; c.7987delGTT<br>p.V2662del" |
| **Validation controls B/LB (#)** | 0; c.1744T>C(p.F582L) and c.2119T>C (p.S707P') (not clear if tested in this assay/data not shown) | 0 | 0 |
| **Statistical analysis (general description)** | uncertain/not specified | uncertain/not specified | uncertain/not specified |
| **Threshold for normal readout** | SF >36% (50.1±13.5%) is considered as radio-normal (Sun et al., 2002). | authors state ~ 1 ICA (<1.5 would work) | not specified, propose % survival <10 at 4 days |
| **Threshold for abnormal readout** | A SF of <21% (13.1±7.2%) has been determined to be the radiosensitive range (Sun et al., 2002). | 2.98–3.20 ICAs per metaphase | not specified, propose % survival >10 at 4 days |
| **Approved assay (y/n)** | Y |  |  |
| **Proposed strength** | **PS3_moderate** (can only be applied if PS3_supporting from kinase assay is met)/<br>**BS3_Supporting** (can be added with BS3_supporting from kinase assay)<br>**No weight** should be applied if only radiosensitivity is available<br>**No weight** should be applied if radiosensitivity and kinase assay are conflicting | **PS3_moderate** (can only be applied if PS3_supporting from kinase assay is met)/<br>**BS3_Supporting** (can be added with BS3_supporting from kinase assay)<br>**No weight** should be applied if only radiosensitivity is available<br>**No weight** should be applied if radiosensitivity and kinase assay are conflicting | **PS3_moderate** (can only be applied if PS3_supporting from kinase assay is met)/ **BS3_Supporting** (can be added with BS3_supporting from kinase assay)<br>**No weight** should be applied if only radiosensitivity is available<br>**No weight** should be applied if radiosensitivity and kinase assay are conflicting |

Note: RNA functional studies reflecting aberrant splicing should be coded as PVS1(RNA) and lack of aberrant splicing as BP7(RNA). Because PS3/BS3 eligible observations measure effects downstream of splicing, it may be appropriate to apply these codes in conjunction with PVS1(RNA)/BP7(RNA).

### Phenotype-related rules (PS4, PM3 and BP2)

#### PS4

Case-control studies with *ATM* pathogenic variants are expected to yield odds ratio (OR) >2 based on the known increased lifetime breast cancer risks for pathogenic variant carriers ^10,11^. ORs should be statistically significant with a p-value <0.05 and a lower 95% confidence interval >1.5.

#### PM3

Biallelic pathogenic variants in *ATM* cause A-T. Laboratory studies are available to help rule out differential diagnoses of other ataxia-associated diseases. Of note, A-T can manifest in an atypical fashion, often called variant A-T, that usually presents in childhood with similar features but has a slower progression. The VCEP has created criteria for patients to meet a ‘confident’ or ‘consistent’ *ATM*-associated A-T phenotype with additional weight afforded to those with a ‘confident’ phenotype. There are several considerations in addition to phenotype that need to be reviewed when weighting and applying PM3, including identification of a second *ATM* variant, phase of the second variant, or zygosity, and general population frequency of the variant under consideration. For the application of PM3, points ascribed to multiple probands are additive and the cumulative points can be used as in Table 5 to assign a final weight.

**TABLE 5.**
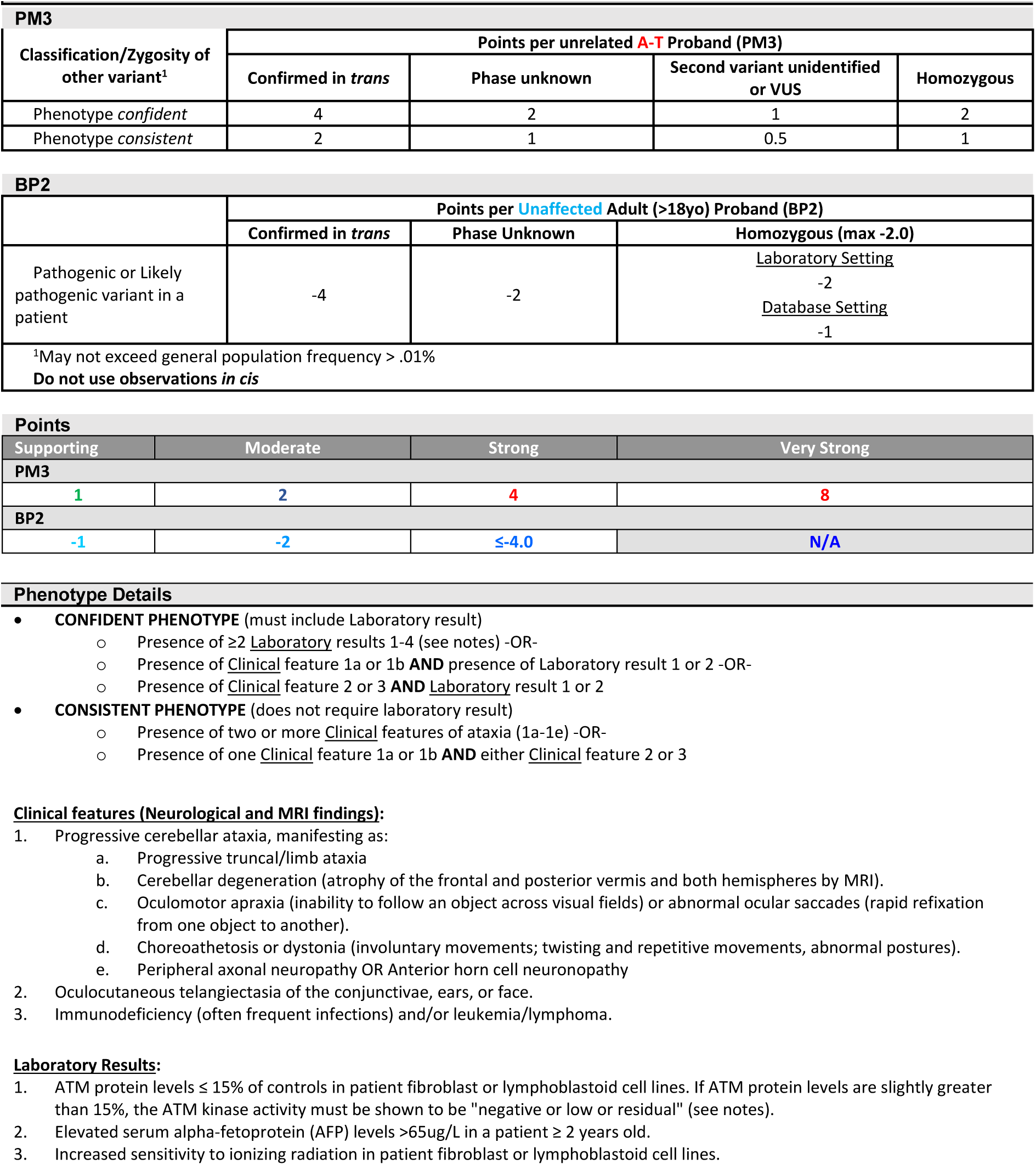
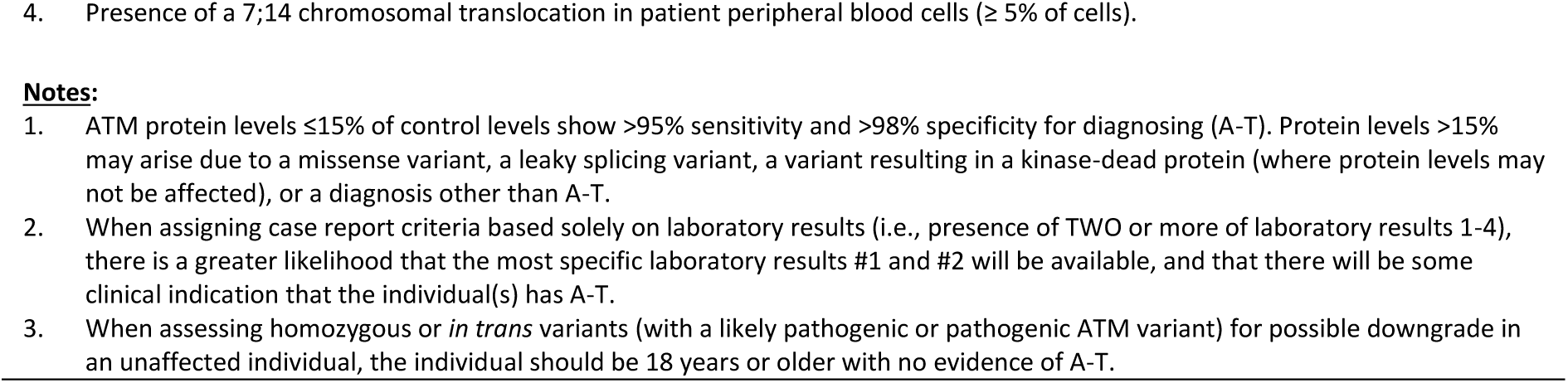
PM3 AND BP2 BIALLELIC CODE STRENGTHS.

#### BP2

Each adult (over 18 years of age) without features of A-T that has an *ATM* variant under consideration in the homozygous state, *in trans,* or phase unknown with a LP/P *ATM* variant contributes to BP2 evidence. There are two important considerations in the application of BP2: 1) The source of the data, where a laboratory setting gets stronger weight than a database setting, due increased rigor in the former and risk of a false positive result due to technical issues like allele drop-out in the latter; and 2) Homozygous individuals have a maximum total weight of −2 points (equivalent to BP2_Moderate) no matter how many independent instances there are. This protects against the assumption that a variant is benign when in reality it might be hypomorphic and pathogenic, but an individual may have sub-clinical or very mild features that may be overlooked by a cancer ascertainment bias. The risk of such a phenomenon is reduced in a compound heterozygous state where the other allele is more likely to have typical risks and stronger presentation. One example of very mild homozygous A-T patients (who are affected with dystonia and not cancer) is caused by the founder alteration *ATM* c.6200C>A (p.Ala2067Asp) ^73^. Excepting homozygous cases, multiple cases of biallelic adult patients unaffected by A-T are additive and can be ascribed BP2 weight based on the cumulative points defined in Table 5 up to a maximum weight of BP2_Strong.

##### Modified Evidence Code Combinations

The HBOP VCEP adopted the original ACMG-AMP categorical evidence code combinations^1^ with two modifications. To achieve a minimum likely pathogenic classification for PVS1-eligible alterations, the combination of PVS1 plus one additional supporting line of pathogenic evidence is allowed to achieve likely pathogenic. In addition, one strong line of evidence in the benign direction is sufficient to achieve a likely benign classification. Both specific modifications are in line with a Bayesian model of variant interpretation published by the SVI^16^ (Table 6). The use of several code combinations is explicitly permitted or restricted by this VCEP and/or the SVI and these are listed in Table 3.

**TABLE 6.** RULES FOR COMBINING CRITERIA.

| TABLE 6. RULES FOR COMBINING CRITERIA |  |
| --- | --- |
| PATHOGENIC CRITERIA |  |
| **Pathogenic** | 1. 1 Very Strong (PVS1, PVS1(RNA) PM3_VeryStrong) AND<br>a. $\geq 1$ Strong (PS1-PS4, PM3_Strong, PP1_Strong) OR<br>b. $\geq 2$ Moderate (PM1-PM6, PP4_Moderate, PP1_Moderate) OR<br>c. 1 Moderate (PM1-PM6, PP4_Moderate, PP1_Moderate) and 1 Supporting (PP1-PP5, PM3_Supporting) OR<br>d. $\geq 2$ Supporting (PP1-PP5, PM3_Supporting)<br>2. $\geq 2$ Strong (PS1-PS4, PM3_Strong, PP1_Strong) OR<br>3. 1 Strong (PS1-PS4, PM3_Strong, PP1_Strong) AND<br>a. $\geq 3$ Moderate (PM1-PM6, PP4_Moderate, PP1_Moderate) OR<br>b. 2 Moderate (PM1-PM6, PP4_Moderate, PP1_Moderate) AND $\geq 2$ Supporting (PP1-PP5, PM3_Supporting) OR<br>c. 1 Moderate (PM1-PM6, PP4_Moderate, PP1_Moderate) AND $\geq 4$ Supporting (PP1-PP5, PM3_Supporting)<br>4. 1 Moderate (PM1-PM6, PP4_Moderate, PP1_Moderate) AND $\geq 4$ Supporting (PP1-PP5, PM3_Supporting) |
| **Likely Pathogenic** | 1. 1 Very Strong (PM3_VeryStrong) AND 1 Moderate (PP1-PP5, PM3_Supporting) OR<br>2. 1 Very Strong (PVS1, PM3_VeryStrong) AND 1 Supporting (PP1-PP5, PM3_Supporting) OR<br>3. 1 Strong (PS1-PS4, PM3_Strong, PP1_Strong) AND 1-2 Moderate (PM1-PM6, PP4_Moderate, PP1_Moderate) OR<br>4. 1 Strong (PS1-PS4, PM3_Strong, PP1_Strong) AND $\geq 2$ Supporting (PP1-PP5, PM3_Supporting) OR<br>5. $\geq 3$ Moderate (PM1-PM6, PP4_Moderate, PP1_Moderate) OR<br>6. 2 Moderate (PM1-PM6, PP4_Moderate, PP1_Moderate) AND $\geq 2$ Supporting (PP1-PP5, PM3_Supporting) OR<br>7. 1 Moderate (PM1-PM6, PP4_Moderate, PP1_Moderate) AND $\geq 4$ Supporting (PP1-PP5, PM3_Supporting) |
| BENIGN CRITERIA |  |
| **Benign** | 1. 1 Stand-Alone (BA1) OR<br>2. $\geq 2$ Strong (BS1-BS4) |
| **Likely Benign** | 1. 1 Strong OR<br>2. 1 Strong (BS1-BS4) and 1 Supporting (BP1-BP7, BS3_Supporting, BP7_Supporting(RNA)) OR<br>3. $\geq 2$ Supporting (BP1-BP7, BS3_Supporting, BP7_Supporting(RNA)) |

### Pilot

Biocurators evaluated 33 variants of varying type and ClinVar classification in a pilot study. Clinical data were collected regarding co-occurrence data from participating clinical diagnostic laboratories and disseminated in a deidentified fashion to the biocurators. Each variant was reviewed independently by two biocurators who applied lines of evidence for a final classification. Evidence codes and classifications were compared among the biocurator group and reviewed by the HBOP VCEP. Evidence codes and classifications approved by the VCEP were submitted for SVI approval and ultimately deposited to ClinVar. The pilot curation set consisted of 10 non-splicing PVS1-eligible alterations (of a variety of variant types); 13 missense alterations (including one generated by an indel); 7 intronic variants; and 3 synonymous variants. Of these 9 had a consensus B/LB classification in ClinVar, 13 had a consensus P/LP classification in ClinVar; 6 had conflicting interpretations, and 5 were considered a VUS. After developing and applying the VCEP rules, the final classifications achieved were 9 benign variants, 2 likely benign variants, 4 likely pathogenic variants, 12 pathogenic variants, and 6 variants of uncertain significance (Figure 3, Table 7).

**Figure 3.**
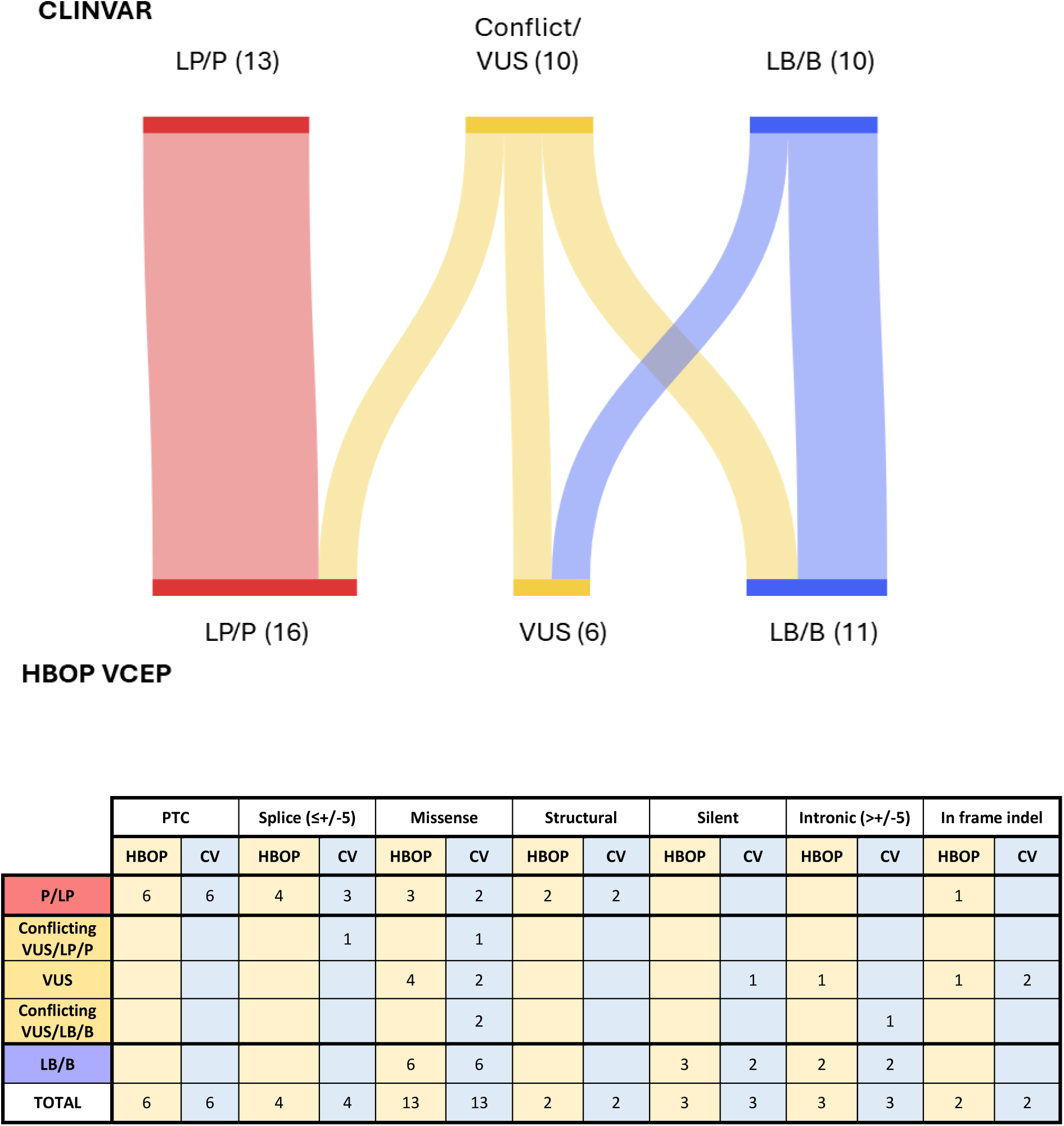
ATM Pilot Variant Categorization. 33 pilot variants are displayed as community classification in ClinVar (left) where VUS/LP/P conflicting interpretation variants and VUS/LB/B conflicting interpretation variants are binned along with consensus VUS as “ClinVar VUS/Conflict”. Interpretation with the HBOP Rules specifications for ATM are on the right. Granular detail of the type of conflict and the type of variant are presented in the table. PTC: Premature Termination Codon; CV: ClinVar

**TABLE 7.**
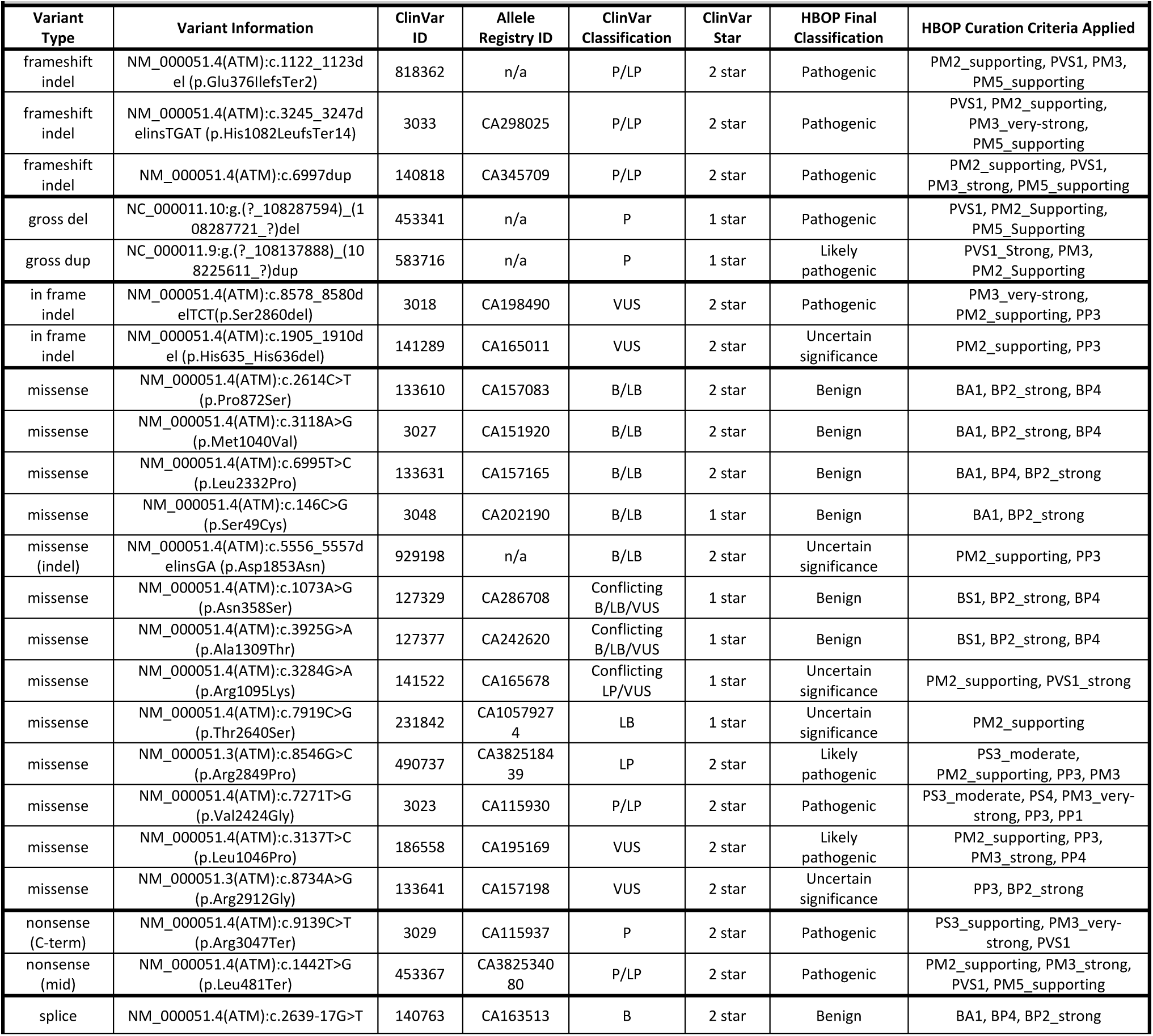
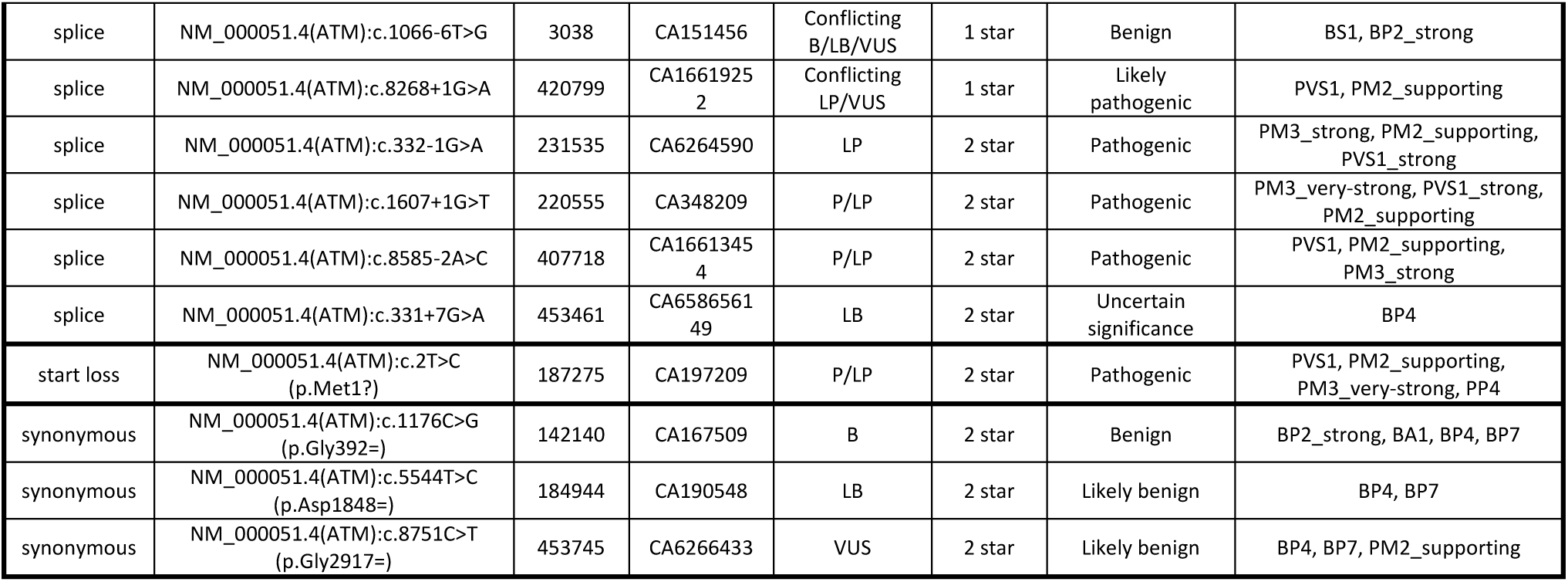
PILOT VARIANTS RESULTS.

Among the variants considered (likely) benign in ClinVar (n=9), the VCEP classified six as (likely) benign and three as VUS (*ATM* c.5556_5557delinsGA (p.Asp1853Asn), *ATM* c.7919C>G (p.Thr2640Ser) and *ATM* c.331+7G>A). Among the variants that were (likely) pathogenic in ClinVar (n=13), the VCEP classified all 13 as (likely) pathogenic. Among 11 variants classified as VUS or conflicting in ClinVar, the VCEP classified five as (likely) benign (four due to application of BA1 or BS1, and one due to the combination of BP4 and BP7), three as (likely) pathogenic (two with PM3_Strong or PM3_Very Strong; and one with the application of PVS1), and three as VUS due to limited evidence (Figure 3, Table 7). The final classifications asserted by the VCEP were submitted to the ClinGen VCI and deposited in ClinVar.

## Discussion

The routine employment of Next-Generation Sequencing represents major advancement in the detection of pathogenic variants in hereditary cancer genes. However, a concomitant and seemingly exponential increase in the detection of variants of uncertain significance is an unfortunate discomfort for many patients and care providers. While it is not possible to resolve the classification of all variants, the development of a set of rules to harmonize classifications across diagnostic and research laboratories can decrease uncertainty related to differential classifications within the public domain. The HBOP VCEP was tasked to define such ACMG/AMP guidelines for *ATM* under the FDA-approved ClinGen VCEP process. This body of work describes the decisions made by the VCEP towards that goal with the ultimate benefit of improving patient outcomes.

The Spanish ATM Working Group (SpATM-WG) defined gene-specific ACMG/AMP style rules for *ATM,* with many similar decisions on rules specifications (Supplementary Table 1)^74^. However, this VCEP also has substantial departures from the SpATM-WG rules that result largely from a more in-depth analysis related to the FDA-approved process that requires ClinGen SVI and HC-CDWG oversight and collaboration related to rules development. For example, this VCEP has justified the up-weighting of the PM3 and BP2 biallelic codes while the SpATM-WG adopted the original SVI-expounded recommendations (https://clinicalgenome.org/site/assets/files/3717/svi_proposal_for_pm3_criterion_-_version_1.pdf). Another difference is the SpATM-WG assignment of PS3 to variants identified in A-T patients who do not have sufficient ATM expression or substrate phosphorylation. The HBOP VCEP considers this a phenotypic line of evidence rather than a functional line of evidence as this result is not necessarily variant-specific, rather a molecular confirmation of the disease-state of the patient. This concept is incorporated into the VCEP interpretation for PM3. Lastly, among other differences, the HBOP VCEP has elected to omit certain codes for A-T patients that SpATM-WG does apply including *de novo* codes PM6 and PS2, co-segregation codes BS4 and PP1, and PS4 proband counting, which this VCEP applies as PM3.

The careful in-depth consideration of each rule has had an impact on ClinVar classified variants leading to a substantial decrease in the conflicting/VUS rate by nearly 50% (ClinVar n=11; VCEP n=6). The improvement of this VUS rate is likely related to three major features: 1) data sharing of otherwise siloed clinical data among participating clinical diagnostic laboratories towards the application of PM3 and BP2 biallelic codes; 2) the establishment of BA1 and BS1 frequency thresholds leading to the increased number of LB/B variants; and 3) the justified increase in weight applied to A-T patients under PM3 leading to the increased number of LP/P variants. The VCEP is performing ongoing curation and further rule modifications taking into consideration any new information that may be forthcoming, including the development of any new ATM functional studies. Using this method, this VCEP aims to further reduce VUS rates and discordance in variant interpretations submitted to ClinVar with the ultimate goal of improving risk assessment and family genetic counseling.

## Data Availability

All data produced in the present work are contained in the manuscript

## ACKNOWLEDGEMENTS

Lidia Feliubadaló, Patrick Concannon, R. Malcolm Taylor, Melissa Southey, Kelly McGoldrick, Sarah Brnich, Sarah Nielson, Huma Rana, Deborah Ritter, Steven Harrison, Sharon Plon, Heidi Rehm, Leslie Biesecker, ClinGen SVI, Hereditary Cancer Clinical domain WG executive committee

---

**Supplementary Figure 1.**
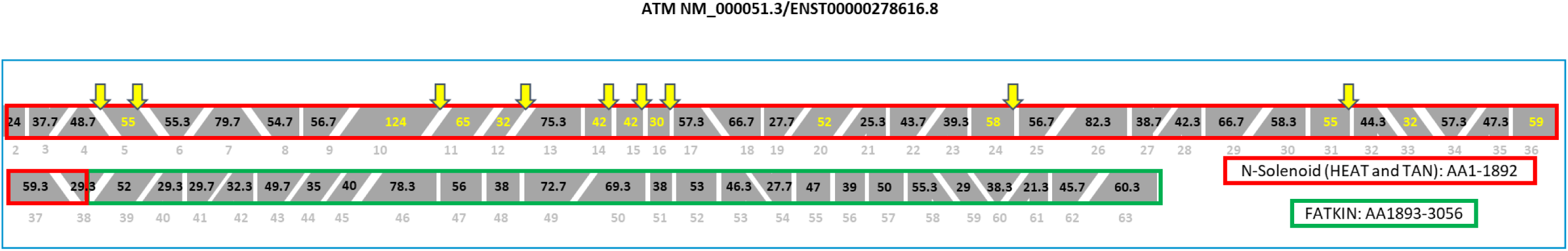
In-Frame Losses in the N-Solenoid Domain Cause A-T. A) The ATM gene depicted exon-by-exon as in Figure 2 (gray text on bottom is total exon numbering) with in frame exons within the N-Solenoid domain indicated by yellow text. Yellow down-facing arrows indicate a variant identified at that splice junction in a patient with A-T. Details of the observed in-frame splice defect, patient genotype and citation are described in the table.

## Supplementary Table 1

**Supplementary Table 1.** N-Solenoid losses for A-T Patients.

| Site c. (Exon) | Observed transcript (p.) | # AA lost | Ataxia-Telangiectasia Patients |
| --- | --- | --- | --- |
| c.332 (Exon 5) | r.332_496del (p.Arg111_Glu166delinsK)<sup>1</sup> | 56 | ATM c.332-1G>A CH with ATM c.2250G>A<sup>1</sup> |
| c.496 (Exon 5) | r.332_496del (p.Arg111_Glu166delinsK)<sup>2</sup> | 56 | ATM c.496+5G>A CH with ATM c.7875_7876delTGinsGC<sup>2-4</sup> |
| c.1607 (Exon 10) | r.1407_1607del (p.Arg469_Cys536delinsSer)<sup>7</sup> | 67 | ATM c.1607+1G>T homozygous and CH with ATM c.3576G>A (p.K1192K) or variant not identified<sup>5-8</sup> |
|  | Intron retention also reported<sup>5</sup> |  |  |
| c.1898 (Exon 12) | r.1803_1898del (p.Asn602_Cys633del)<sup>9</sup> | 32 | ATM c.1898+1G>T CH with ATM EX21_29del<sup>9</sup>; ATM c.1898+2T>G CH with ATM c.5825C>T (p.A1942V), c.3085_6086insA, or unidentified 2nd variant<sup>10-12</sup> |
| c.2250 (Exon 14) | r.2125_2250del (p.Ile709_Lys750del) (Ambry, internal data) | 42 | ATM c.332-1G>A CH with ATM c.2250G>A<sup>1</sup> |
| c.2376 (Exon 15) | r.2251_2376del (p.Ser751_Lys792del) (Ambry, internal data) | 42 | ATM c.2376+1G>A CH with ATM c.7875_7876delTGinsGC<sup>13</sup> |
| c.2466 (Exon 16) | r.2377_2466del (p.Lys793_Leu822del) (Ambry, internal data) | 30 | ATM c.2466+1delG CH with ATM c.6913C>T (p.Q2305*) or unidentified 2nd variant<sup>5, 9, 14</sup> |
| c.3576 (Exon 24) | r.3403_3576del (p.Ser1135_K1192del)<sup>17,18</sup> | 58 | ATM c.3576G>A CH with c.3111delT, c.2413C>T (p.Arg805*), c.1369C>T (p.Arg457*), c.4842_4843insCT, c.8545C>T (p.Arg2849*); c.1607+1G>T; c.6761A>C (p.His2254Pro))<sup>5,9,15,16,17</sup>; ATM c.3576G>A homozygous (2 patients)<sup>7,13,15,18</sup> |
| c.4776 (Exon 31) | r.4612_4776del165 (p.V1538_E1592del)<sup>20</sup> | 55 | ATM c.4776+1G>T CH with ATM c.748C>T (p.Arg250*); c.8977C>T (p.Arg2993*); or unknown variant<sup>11,19,20</sup>; ATM c.4776+2T>C homozygote<sup>5,7</sup> |
| CH = Compound Heterozygous |  |  |  |

---

**Supplementary Table 2.**
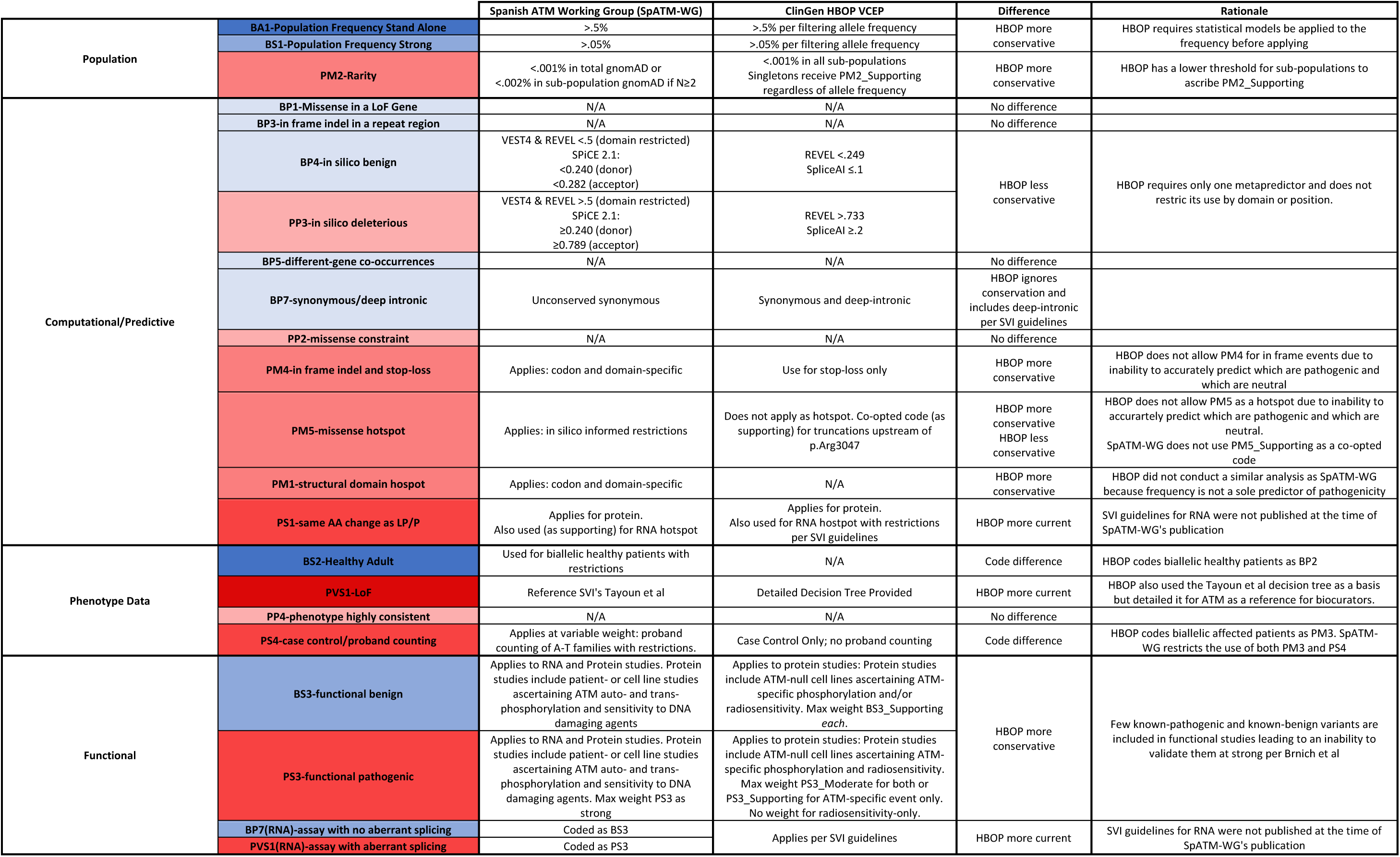
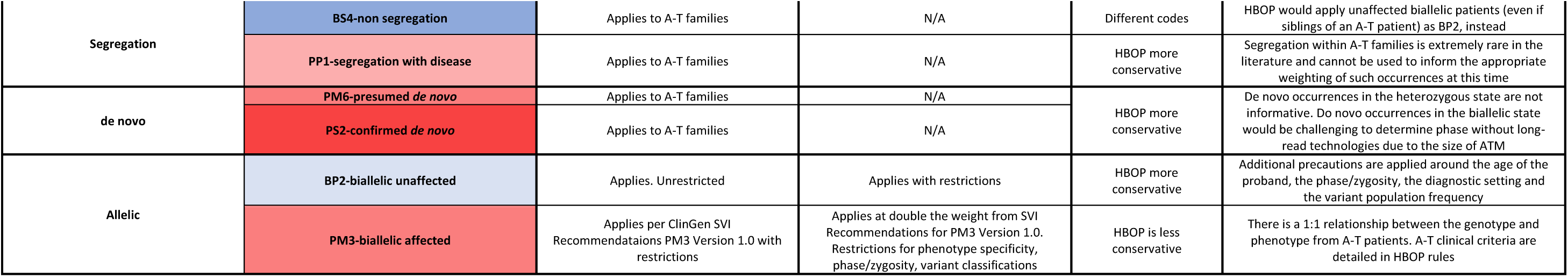
SpATM-WG vs HBOP Classification Criteria Comparison.

